# Increased rostral medial frontal GABA+ in early psychosis is obscured by levels of negative affect

**DOI:** 10.1101/2022.08.30.22279365

**Authors:** Molly Simmonite, Beier Yao, Robert C. Welsh, Stephan F. Taylor

**Author notes:** Corresponding author: Molly Simmonite, Department of Psychiatry, Rachel Upjohn Building, 4350 Plymouth Road, Ann Arbor, MI, 48109.

## Abstract

**Background:** Evidence suggests dysfunction of GABAergic interneurons in psychosis, and prior research has linked GABAergic function with a tendency toward negative affective states. Magnetic resonance spectroscopy (MRS) studies measuring GABA have yielded inconsistent findings. We investigate GABA concentrations in young adults with attenuated psychosis syndrome (APS) and first episode psychosis (FEP), as well as testing the hypothesis that negative affect is a clinical phenotype that is associated with reduced GABA.

**Materials:** MRS data were obtained from 14 patients with FEP, 7 patients with APS and 15 healthy controls (HC), using a MEGA-PRESS sequence on a 3T Philips Ingenia scanner. Voxels were placed in rostral MFC and midline-occipital cortex. Gannet 3.1 was used to determine GABA+ and Glx (glutamate and glutamine combined) concentrations.

**Results:** We found a trend towards increased rostral MFC GABA+ concentrations in FEP, but no group differences in occipital GABA+ concentrations. When covarying for scores on the Psychological Stress Index, rostral MFC GABA+ levels in FEP were significantly greater than APS and HC. Planned comparisons revealed a trend towards increased rostral MFC GABA+ in APS relative to HC. No group differences in Glx or occipital GABA+ were found.

**Conclusion:** These results, considered alongside previously published findings, suggest multiple factors influencing GABA+ in psychosis. We conclude a process exists which drives up GABA+ in early psychosis, alongside a separate process in which reduced GABA+ is associated with increased negative affect. These multiple processes have resulted in contradictory findings, and their untangling is critical to understanding of GABA+ in psychosis.

## 1. INTRODUCTION

Considerable evidence implicates dysfunction of GABAergic systems in schizophrenia, alongside other psychosis spectrum disorders. Post-mortem findings of reductions in the expression of glutamic acid decarboxylase-67 (GAD67), an enzyme critical for GABA synthesis, have been well replicated in both schizophrenia (Curley et al., 2011; Guidotti et al., 2000; Lewis et al., 2012; Schmidt & Mirnics, 2015) and bipolar disorder (Guidotti et al., 2000; Torrey et al., 2005; Woo et al., 2008), and observed in both the cerebral cortex and hippocampus. GAD67 reductions also have been found in rodent models of schizophrenia (Behrens et al., 2007; Liu et al., 2001; Takao et al., 2013). Using a pharmacological challenge with lorazepam, a non-subtype selective benzodiazepine that potentiates GABA function, in combination with blood oxygen-level dependent (BOLD) fMRI, our group has demonstrated GABAergic dysfunction in schizophrenia both while patients passively viewed emotionally salient images (Taylor et al., 2014) and while performing a face processing task (Tso et al., 2015). Further, the altered BOLD response to lorazepam in schizophrenia was positively correlated with increased negative affect.

While traditionally there has been a focus on the association of GABA with cognitive deficits, emerging links between GABAergic systems, affect, and emotion processing indicate an important line of inquiry. Affective dysregulation is often exhibited by patients with psychosis spectrum disorders (Myin-Germeys & van Os, 2007; Phillips et al., 2007). We conceptualise this dysregulation as a high propensity to experience negative affect states such as anxiety and dysthymia, and to exhibit a low tolerance for even minor, everyday stressors. Patients with schizophrenia report more negative experiences in daily life (Docherty, 1996; Horan et al., 2005; Jones & Fernyhough, 2007; Myin-Germeys et al., 2000) and perceive emotional stimuli with a more negative bias in laboratory settings (Burbridge & Barch, 2002; A. S. Cohen & Minor, 2010; Holt et al., 2006). Importantly, measures of negative affect have significant prognostic value in schizophrenia, predicting hospitalisation and poor functional outcome (Blanchard et al., 1998; Grove et al., 2016; Horan et al., 2008).

Here, we aimed to build upon findings from our group, which demonstrated negative affect associated GABAergic abnormalities in schizophrenia using in-vivo functional imaging, by exploring these questions using magnetic resonance spectroscopy. Thus far, studies of GABA concentrations in schizophrenia using magnetic resonance spectroscopy have reported mixed findings, with evidence for increased (de la Fuente-Sandoval et al., 2018; Öngür et al., 2010; Yang et al., 2015), decreased (Goto et al., 2009; Menschikov et al., 2016; Thakkar et al., 2017; Yoon et al., 2010) and as normal GABA concentrations when compared with healthy controls (C. M. A. Chen et al., 2014; Tayoshi et al., 2010). These divergent findings likely reflect the influence or multiple factors or processes upon GABAergic systems, such as illness stage and symptom profile, as well as the location of the MRS voxel in the brain. Indeed, when examining rostral medial frontal cortex (MFC) GABA concentrations, several groups studying early, unmedicated patients have revealed elevated GABA concentrations (T. Chen et al., 2017; de la Fuente-Sandoval et al., 2018; Kegeles et al., 2012), although not all groups have replicated this finding (e.g. Wang et al., 2016). By contrast, studies of older medicated patients have revealed no abnormalities (Chiu et al., 2018; Kegeles et al., 2012; Rowland et al., 2013), or reductions when compared with healthy controls (Rowland et al., 2013; Thakkar et al., 2017).

In this study, we used MRS to compare GABA+ concentrations in the rostral MFC and midline-occipital cortex in patients with first episode psychosis (FEP) and individuals meeting criteria for attenuated psychosis syndrome (APS) with healthy, age-matched controls. Additionally, we explore links between GABA+ concentrations and levels of negative affect (NA) as measured by the Psychological Stress Index (PSI9: Tso et al., 2012). Based on prior reports of increased rostral MFC GABA in early schizophrenia, we hypothesized that GABA+ concentrations in this voxel would be increased relative to controls. We also predicted, based on findings linking increased NA with altered BOLD responses, that GABA+ concentrations would be associated with NA. As glutamate has been linked with emotion processing in healthy controls (Stan et al., 2014), as well as reported abnormalities in the excitation/inhibition balance in psychosis (for a review see Liu et al., 2021), we also performed an exploratory analysis examining relationships between NA and Glx (the combined resonance of glutamate and glutamine).

## 2. METHODS

### 2.1 Participants

Study participants were 14 patients with FEP, 7 individuals meeting APS criteria, and 15 healthy controls (HC) who were group matched to the psychosis groups in terms of age, gender, and years of parental education (Table 1). Participants were recruited from the local community via advertisements and referrals from clinicians at local mental health clinics. The Structured Interview for Psychosis Syndromes (SIPS: Miller et al., 2003) and Structured Clinical Interview for DSM-IV Axis I Disorders (SCID-IV: First et al., 1996) were used to determine APS status and psychiatric diagnoses. Participants completed the 9-item PSI9 (Tso et al., 2012), a validated self-report scale which was modelled on Perceived Stress Scale (S. Cohen et al., 1983). The PSI9 was designed to measure negative affect and stress sensitivity over the last month, but can also capture trait-like tendencies, and has demonstrated predictive power for both functional and clinical outcomes (Tso et al., 2012).

**Table 1:** Participant demographics and clinical characteristics

| Table 1: Participant demographics and clinical characteristics |  |  |  |  |
| --- | --- | --- | --- | --- |
|  | HC<br>(n = 15) | APS<br>(n = 7) | FEP<br>(n = 14) | Group<br>differences |
| *Demographics* |  |  |  |  |
| Age | 21.60 (±3.56) | 19.86 (±3.63) | 21.64 ±2.56 | $F_{2,33} = 0.85, p = .44$ |
| Gender |  |  |  |  |
| Men | 10 (66.67%) | 5 (71.43%) | 9 (64.29%) | $\chi^2(2, N = 36) = 0.11, p = .95$ |
| Women | 5 (33.33%) | 2 (28.57%) | 5 (35.71%) |  |
| Handedness<sup>a</sup> | 35.01 (±58.80) | 62.27 (±14.15) | 53.29 (±61.00) | $F_{2,33} = 0.73, p = .49$ |
| Socioeconomic status | 2.20 (±0.41) | 2.29 (±0.76) | 2.14 (±0.53) | $\chi^2(6, N = 36) = 7.31, p = .29$ |
| Parental education (years) | 16.27 (±1.98) | 16.14 (±2.04) | 17.00 (±3.59) | $F_{2,33} = 0.35, p = .71$ |
| Duration of illness (years)<sup>b</sup> | - | 4.31 (±2.76) | 1.44 (±1.03) |  |
| *Medications* |  |  |  |  |
| CPZeq (mg daily) | - | 133.33* | 238.10 (±223.27) |  |
| *Clinical assessments* |  |  |  |  |
| PANSS positive | - | 13.71 (±1.70) | 15.14 (±4.62) | $t_{18.13} = -1.03, p = .32$ |
| PANSS negative | - | 10.00 (±1.91) | 16.43 (±6.74) | $t_{16.60} = -3.31, p = .004$ |
| PANSS general psychopathology | - | 23.29 (±2.69) | 29.29 (±6.01) | $t_{18.92} = -3.16, p = .005$ |
| PANSS total | - | 47.00 (±3.56) | 60.86 (±11.47) | $t_{17.11} = -4.14, p < .001$ |
| *Functional assessments* |  |  |  |  |
| PSI9 | 9.87 ± 4.09 | 15.14 (±6.07) | 19.29 (±5.70) | $F_{2,33} = 12.14, p < .001$ |
| Note: CPZeq = antipsychotic dose in chlorpromazine equivalent mg daily; PANSS = positive and Negative Symptoms Scale; PSI9 = 9-item Psychological Stress Index. *only 1 member of the APS group was receiving medication, and their dosage is reported here.<br><sup>a</sup>assessed using the Edinburgh Handedness Inventory<br><sup>b</sup>for the FEP group, durations are calculated as time from the date psychosis was developed, to participation in the study. For APS participants, durations were calculated from the estimated onset of subclinical psychosis symptoms. For APS participants to be eligible for inclusion in the study, their symptoms had to have worsened in the past year. |  |  |  |  |

All participants were aged 15 – 27 years old, with normal, or corrected to normal vision. Exclusion criteria included contraindications to MRI (e.g., pacemakers, metallic implants), alcohol or substance use in the past month or dependence in the last 6 months, history of head injury or concussions with unconsciousness lasting > 5 minutes. Healthy control participants were free from histories of psychiatric disorders, and history of psychotic or bipolar disorder in their first-degree relatives.

Study procedures were approved by the Institutional Review Board of the University of Michigan Medical School. Following full explanation of the study, written informed consent was obtained from participants aged 18 years old and over. For those under 18 years, written parental consent was obtained, alongside written assent from the participant.

### 2.2 MR acquisition

MR data were acquired using a 3T Phillips Ingenia system with a 32-channel head coil. Head motion was minimized using padding and Velcro straps. For anatomical reference and tissue classification, we obtained a structural scan for each participant (3D-GRE, slice thickness = 1.4 mm, TR = 25 ms, TE = minimum, flip angle = 35, field of view = 24 cm).

For the assessment of GABA+ and Glx, MRS data were collected using a MEGA-PRESS sequence (Mescher et al., 1998), with the following parameters: TE = 68 ms (TE1 = 15 ms, TE2 = 53 ms); TR = 1.8 seconds; 256 transients of 2000 data points; spectral width 2 kHz; frequency selective editing pulses (14 ms) applied at 1.9 ppm (ON) and 7.46 ppm (OFF).

Data were collected from two voxels (Figure 1): a 34 × 36 × 20mm rostral MFC voxel placed anterior to the genu of the corpus collosum, and a 35 × 25 × 25mm occipital cortex voxel centered on the calcarine sulcus, parallel to the cerebellar tentorium. Acquisition times were 11 mins 24 sec for the MFC voxel and 11 mins 12 sec for the occipital voxel.

**Figure 1:**
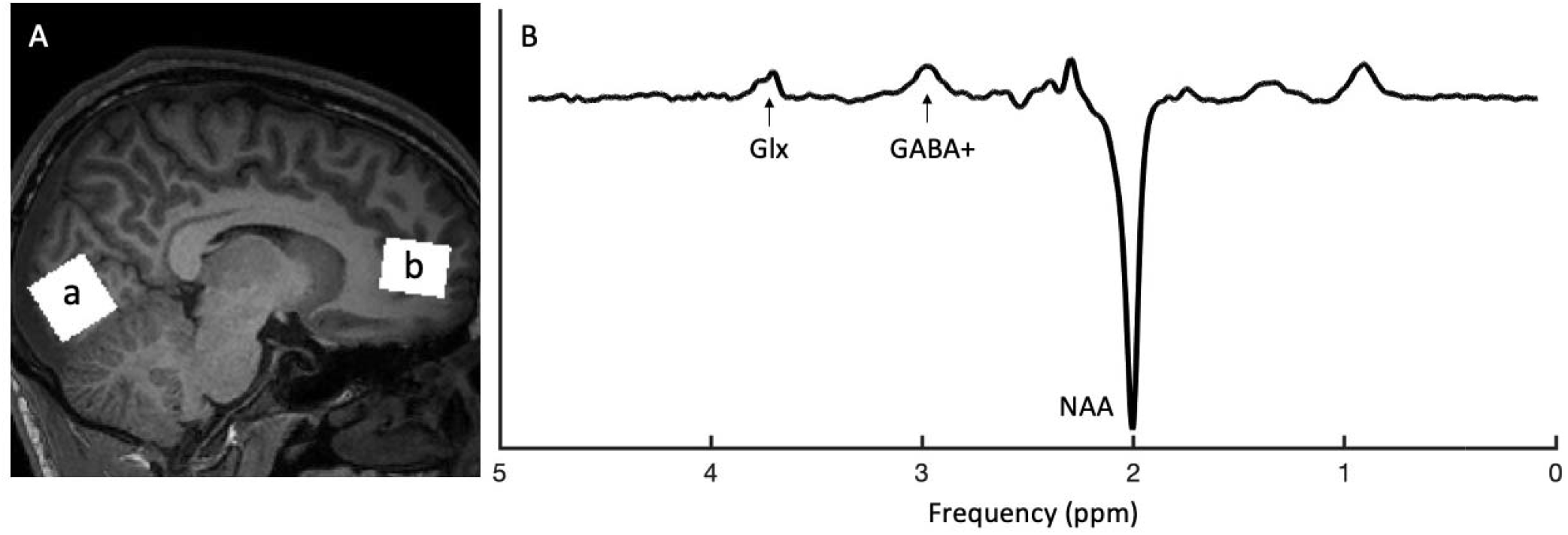
(A) Example MRS voxels placements). Voxel a illustrates placement of the midline occipital voxel and voxel b illustrates the rostral medial frontal voxel. (B) Example MEGA-PRESS spectra from one participant, with Glx, GABA and NAA peaks labeled.

Recommended minimum reporting details for the MRS data are included in Supplemental Table 3, set out as per Minimum Reporting Standard for in-vivo Magnetic Resonance Spectroscopy (MRSinMRS): Experts’ Consensus Recommendations (Lin et al., 2021).

### 2.3 Data Preprocessing and Analysis

Preprocessing and quantification of the edited MRS spectra were performed using Gannet 3.1 (Edden et al., 2014), an open-source MATLAB based toolbox. Standard processing steps including frequency and phase corrections of time-resolved data using spectral registration were used. GABA and Glx levels at 3.00 and 3.75 ppm respectively were estimated using nonlinear, least-squares fitting. As the GABA signal detected at 3.00 pm using our experimental parameters is expected to contain contributions from macromolecules and homocarnosine, it is referred to as GABA+ throughout this manuscript.

GABA+ and Glx concentrations were evaluated relative to the unsuppressed water signal. While there is debate regarding the best choice of reference metabolite (Mullins et al., 2014), evidence suggests that Cr is decreased in schizophrenia (Öngür et al., 2009), thus we opted against this as our primary measure. Nevertheless, many studies use Cr as a baseline, and we report GABA+/Cr and Glx/Cr in our supplementary materials, to allow comparison with these reports.

Since psychosis, even in its early stages, is associated with morphological changes in the brain such as frontal and temporo-limbic reductions in grey matter (Takahashi & Suzuki, 2018), we calculated estimates of GABA+ and Glx that are corrected for the tissue composition of the MRS voxel. Specifically, Gannet 3.1 coregisters the MRS voxels on to the T1-weighted structural images which we acquired during the same scanning session, then uses SPM12 segmentation functions to calculate the fractions of grey matter (GM), white matter (WM) and cerebrospinal fluid (CSF) in each voxel. As CSF contains negligible amounts of GABA or Glx, Gannet 3.1 uses these fractions to remove the impact of CSF fraction on the voxel, as well as applying tissue specific relaxation and water visibility constraints for each of the three tissue types (Harris et al., 2015). We present the alpha-correction in the main manuscript as our primary measure, which assumes the concentration of GABA+ and Glx in GM is twice that of WM (Ganji et al., 2014) and applies an alpha of 0.5 to correct for between-subject differences in voxel tissue composition. Uncorrected measures referenced to water and creatine included in the supplementary materials.

Spectra were visually inspected for artefacts across Gannet 3.1’s preprocessing steps and as well as goodness of model fit. Gannet 3.1 provides a measure of fit error and to ensure the robustness of our findings, only estimates of GABA+ and Glx with a fit error of less than 15% were included in our analysis. Gannet 3.1 also provides spectra quality metrics including signal to noise ratio (SNR) and linewidth (FWHM) which were also assessed, and spectra with values greater than 3 SD of the sample mean for these metrics were excluded. Any extreme GABA+ values of greater than 5 SD of the sample mean were excluded, to prevent these values having a high influence on statistical analyses.

### 2.4 Statistical analyses

All statistical analyses were performed in R. Group comparisons of demographic and clinical measures were performed using ANOVA, or chi-square tests where measures were categorical. To investigate group differences in GABA+ and Glx, we performed ANOVA, followed by ANCOVA which included PSI9 scores as a covariate. Since our primary measure of interest was GABA+ concentrations in the two voxels, we controlled for multiple comparisons by setting our significance threshold at 0.025 (representing a Bonferroni correction of 0.05/5). Following significant main effects of group, planned comparisons were used to compare GABA+ concentration differences between the FEP group and the APS and HC participants (FEP vs HC + APS), and between APS and HC.

## 3. RESULTS

### 3.1 Sample characteristics

Summaries of demographic and clinical measures are presented in Table 1. Groups did not differ on age, gender, handedness, parental education, or socioeconomic status. APS and FEP groups did not differ on PANSS positive scores, however the FEP group had higher PANSS total scores (t_17.11_ = -4.14, *p*<.001), as well as higher PANSS negative (t_16.60_ = 3.31, *p* = .004) and PANSS general psychopathology (t_18.92_ = -3.16, *p* = .005) scores. There was a significant group difference in PSI9 scores (F_2,33_, = 12.14, *p*<.001), with post-hoc Tukey tests showing PSI9 scores differed significantly between the FEP and HC groups (*p* < .0001), but not between the APS and HC groups (*p* = .08) or between the FEP and APS groups (*p* = .21).

Figure 2 presents the degree of spatial overlap for both voxels, alongside their edited spectra. In the rostral MFC voxel, GABA+ data was excluded from three FEP participants and one HC due to GABA+ fit errors of greater than 15%. Extreme GABA+ values (greater than 5 SDs from the mean) led to the exclusion of rostral MFC GABA+ and Glx from one FEP participant, and Occ GABA+ and Glx from one APS participant.

**Figure 2:**
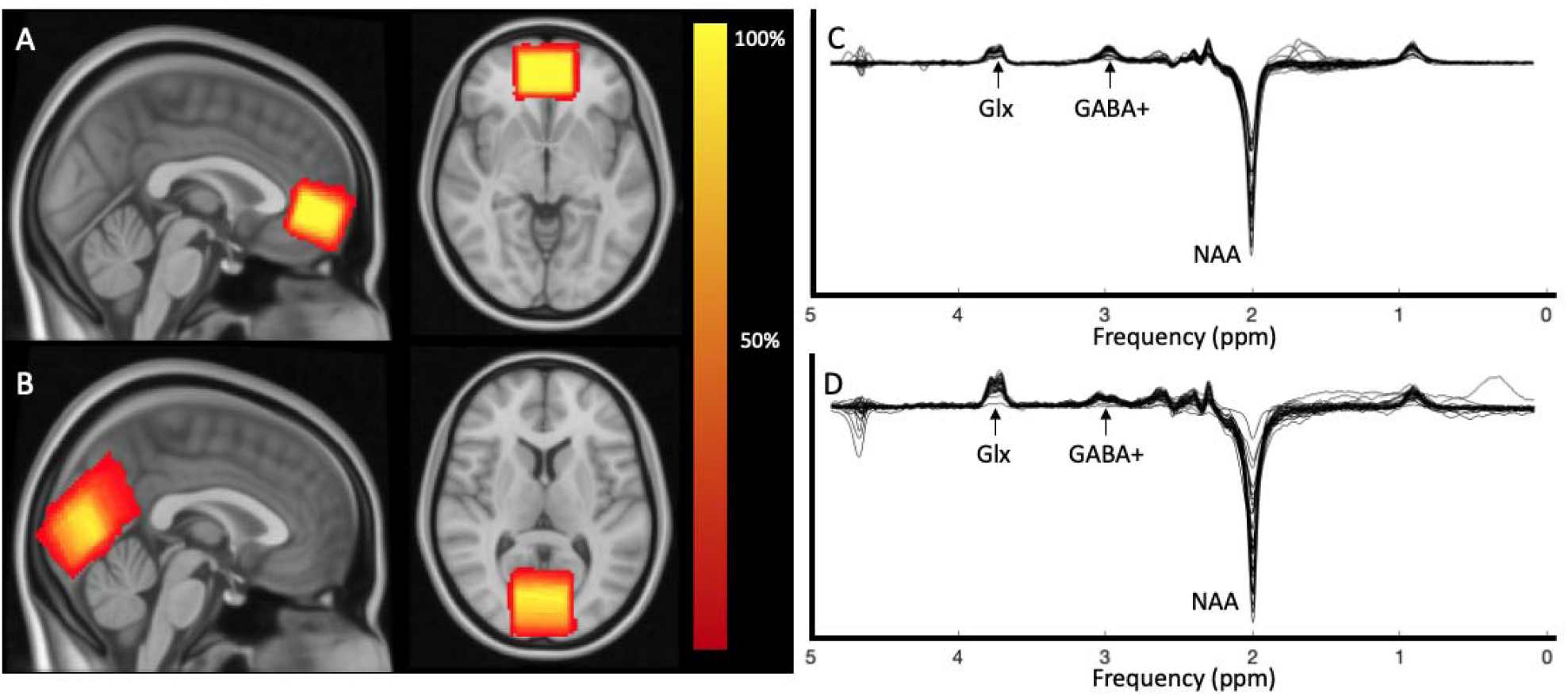
Heat maps describing spatial overlap of MRS voxels in (A) rostral MFC and (B) midline occipital cortex voxel across all participants. Corresponding edited spectral from (C) rostral MFC voxel and (D) midline occipital cortex voxel from all participants (overlaid)

### 3.2 Voxel tissue composition and spectral quality

Table 2 includes a breakdown of voxel tissue compositions. No group tissue composition differences were found in the rostral MFC voxel (GM: F_2,32_ = .92, *p* = .41; WM: F_2,32_ = .96, *p* = .39; CSF: F_2,32_ = 0.20, *p* = .82). In the occipital voxel, there was a significant main effect of group on GM fraction (F_2,32_ = 4.16, *p* = .02), with significantly greater GM fraction in HC participants compared with FEP (p =.01). Group differences in WM fraction trended towards significance (F_2,32_ = 2.59, *p* = .09). There were no group differences in CSF fraction (F_2,32_ = .40, *p* = .68).

**Table 2:** Voxel composition of the rostral MFC and midline occipital cortex voxels

| Voxel | Group | CSF fraction | GM fraction | WM fraction |
| --- | --- | --- | --- | --- |
| Rostral MFC | HC | .14 ($\pm .02$) | .52 ($\pm .04$) | .33 ($\pm .04$) |
| | APS | .14 ($\pm .02$) | .53 ($\pm .03$) | .33 ($\pm .02$) |
| | FEP | .14 ($\pm .04$) | .51 ($\pm .02$) | .35 ($\pm .03$) |
| Occipital | HC | .07 ($\pm .04$) | .60 ($\pm .05$) | .33 ($\pm .06$) |
| | APS | .06 ($\pm .02$) | .59 ($\pm .03$) | .35 ($\pm .02$) |
| | FEP | .07 ($\pm .04$) | .56 ($\pm .03$) | .37 ($\pm .04$) |

GABA+ and Glx fit errors are presented in Table 3. There were no group differences in GABA+ fit error % (MFC: F_2,28_ = 1.57, *p* = .27; Occ: F_2,32_ = 0.22, *p* = .81) or Glx fit error % (MFC: F_2,32_ = .71, *p* = .50; Occ: F_2,32_ = 0.34, *p* = .72). MRS spectral quality measures are presented in Supplemental Table 1. There were no group differences in linewidth (FWHM) or SNR.

**Table 3:** Metabolite concentrations and fit errors (%) in the rostral MFC and midline occipital cortex

| Voxel | Group | GABA+ |  | Glx |  |
| --- | --- | --- | --- | --- | --- |
|  |  | Fit Error (%) | GABA+ | Fit Error (%) | Glx |
| Rostral MFC | HC | 10.96 ($\pm 2.24$) | 2.16 ($\pm 0.42$) | 4.19 ($\pm 0.88$) | 14.69 ($\pm 1.85$) |
| | APS | 11.60 ($\pm 2.62$) | 2.04 ($\pm 0.14$) | 4.08 ($\pm 0.56$) | 14.20 ($\pm 1.67$) |
| | FEP | 9.59 ($\pm 2.62$) | 2.50 ($\pm 0.53$) | 4.77 ($\pm 2.20$) | 14.42 ($\pm 4.03$) |
| Occ | HC | 4.90 ($\pm 2.07$) | 3.24 ($\pm 0.40$) | 4.55 ($\pm 1.29$) | 11.27 ($\pm 1.57$) |
| | APS | 4.91 ($\pm 1.29$) | 3.43 ($\pm 0.51$) | 4.21 ($\pm 2.10$) | 11.11 ($\pm 1.56$) |
| | FEP | 5.39 ($\pm 2.42$) | 3.24 ($\pm 0.41$) | 4.91 ($\pm 1.33$) | 10.72 ($\pm 2.17$) |

### 3.3 Group Differences in GABA+ and Glx

Group average metabolite concentrations are presented in Table 3 and Figure 3. When we conducted ANOVAs to examine group differences in GABA+, we found a trend towards a significant effect of group in the MFC (F_2,28_ = 3.04, *p* = .06), but no differences in the occipital cortex (F_2,32_ = 0.49, *p* = .62). There were no main effects of group on Glx concentration in either voxel (MFC: F_2,28_ = 0.08, *p* = .93; Occ: F_2,32_ = 0.34, *p* = .72).

**Figure 3:**
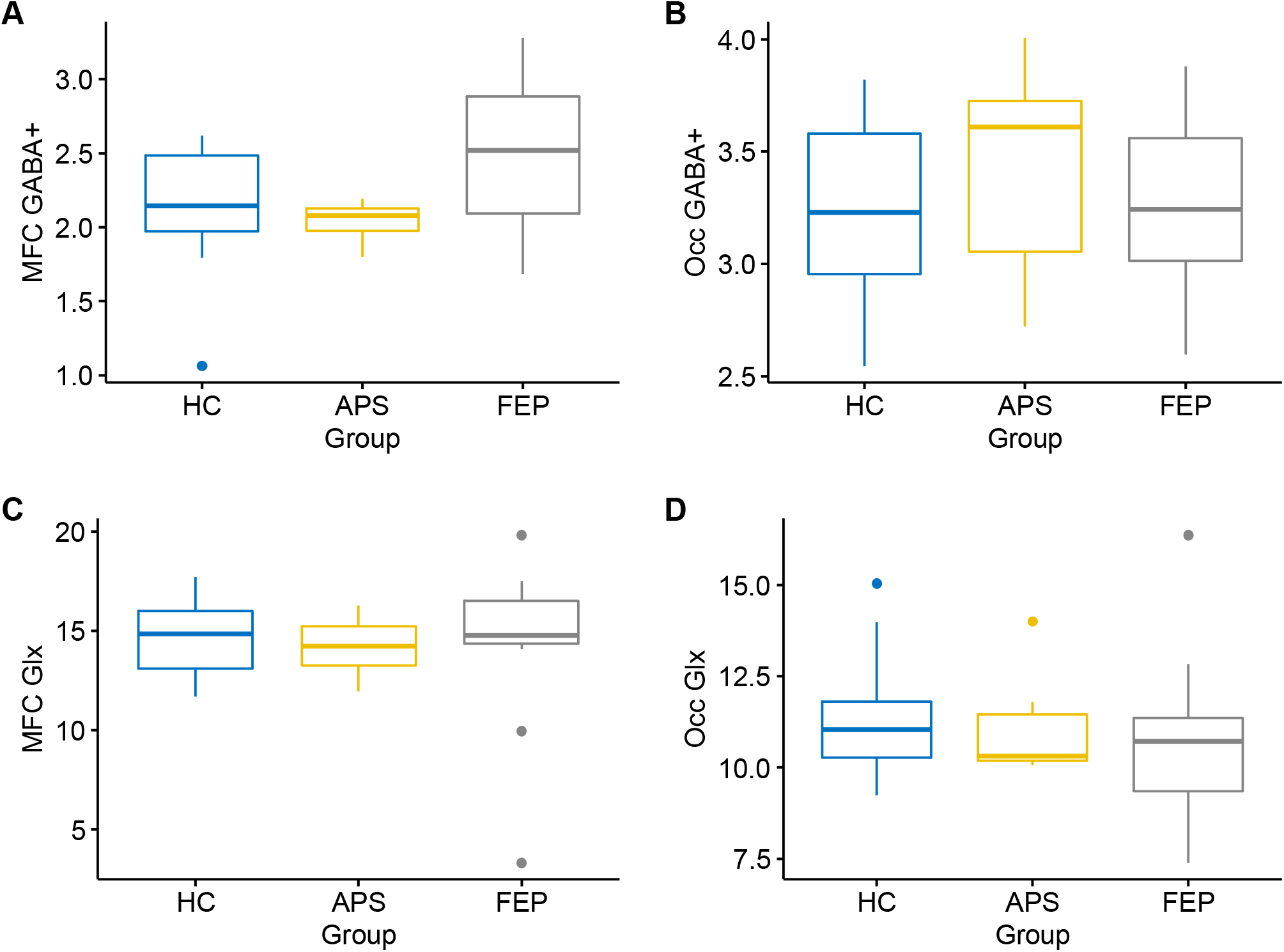
Box plots demonstrating GABA+ concentrations in (A) rostral MFC and (B) midline occipital cortex and Glx concentrations in (C) rostral MFC and (D) midline occipital cortex in each of the three participant groups. HC: healthy controls; APS: attenuated psychosis syndrome, FEP: first episode psychosis. These values are uncorrected for PSI9.

When PSI9 was included in the model to adjust for group differences in negative affect, we found a significant main effect of group on MFC GABA+ concentrations (F_2,28_ = 4.26, *p* = .02). Planned comparisons revealed elevated MFC GABA+ in the FEP group in comparison with the HC and APS groups (t_27_= 2.90, *p* = .01). There was a trend towards elevated MFC GABA+ in the APS group when compared with healthy controls (t_27_= 2.00, p = .0548). We did not find significant effects of group on rostral MFC Glx (F_2,31_ = 0.26, *p* = .77), or occipital GABA+ (F_2,31_ = 0.48, *p* = .62) or Glx (F_2,32_ = 1.03, *p* = .23) when PSI9 was included in the model.

### 3.4 Relationships between GABA+ and anti-psychotic medication dose

In the FEP group, correlations between rostral MFC GABA+ and CPZeq were r = .53, *p* = .11. Correlations between midline occipital GABA+ and CPZeq were r = .07, *p* = .81. Since only one participant with APS was receiving anti-psychotic medication, we did not calculate correlations between GABA+ and CPZeq in this group.

**Figure 4:**
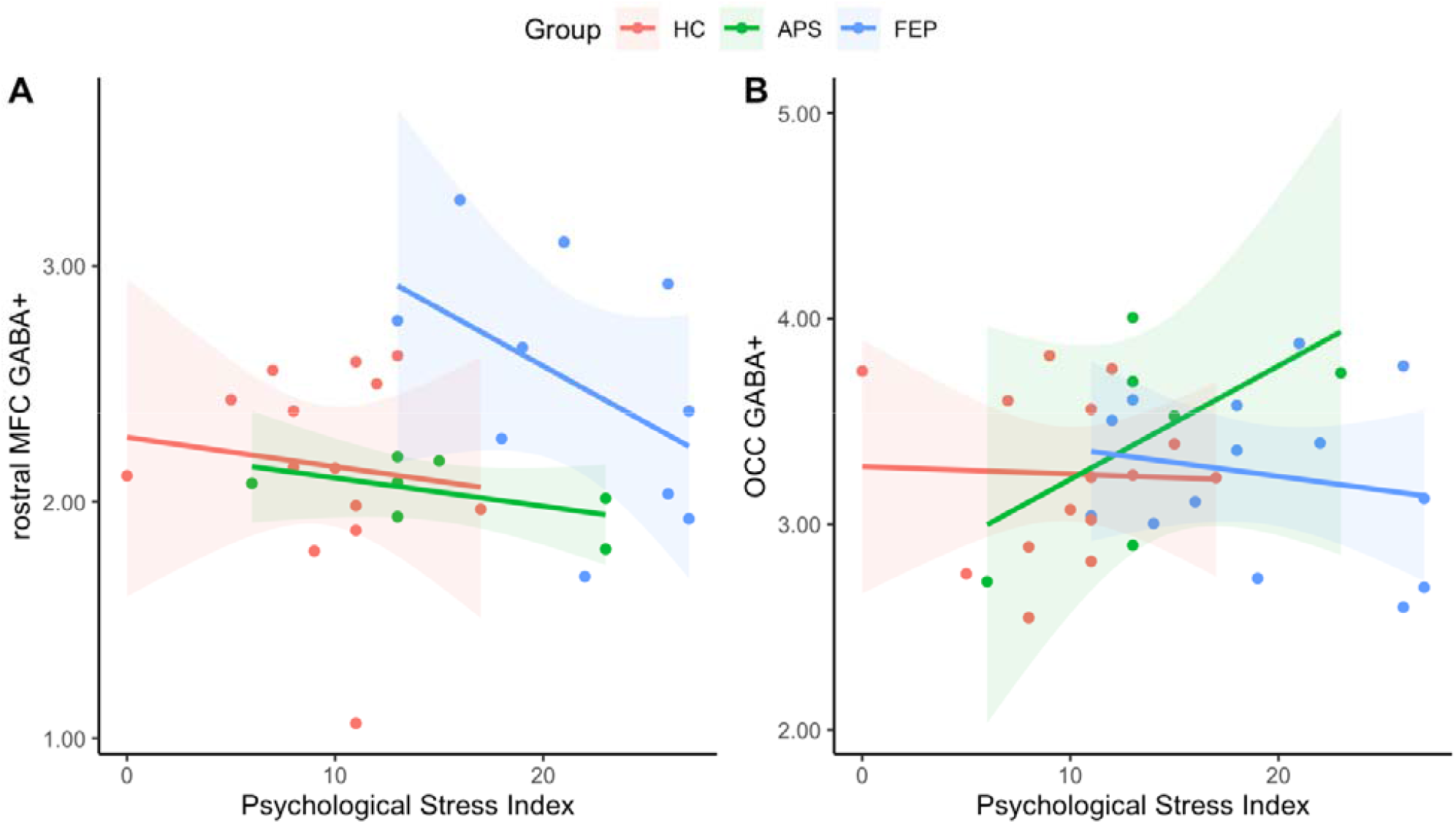
Scatterplots showing relationships between scores on the Psychological Stress Index and GABA+ concentrations in (A) the rostral MFC GABA+ and (B) midline occipital cortex

## 4. DISCUSSION

Using MRS, we investigated rostral MFC and midline occipital cortex GABA+ and Glx concentrations in participants with FEP and APS. With negative affect (NA), measured by PSI9, as a covariate, we found significantly elevated rostral MFC GABA+ in patients with FEP as well as a trend towards elevated rostral MFC GABA+ in individuals meeting APS criteria. This elevation of GABA +concentrations is consistent with prior investigations of rostral GABA in acute schizophrenia, in unmedicated individuals (T. Chen et al., 2017; de la Fuente-Sandoval et al., 2018; Kegeles et al., 2012), although some groups have also reported decreased GABA concentrations in the region of this rostral voxel (Chiu et al., 2018; Wang et al., 2016).

Prior divergent MRS findings and the relatively small number of studies leads us to believe that there are multiple factors influencing GABA levels in psychosis. Synthesizing the results of the current study with previously published reports from our group in which a benzodiazepine challenge was used to demonstrate an abnormal BOLD response in schizophrenia which correlated with levels of negative affect (Taylor et al., 2014; Tso et al., 2015), we suggest there is developing evidence to support at least two separate pathological processes involving rostral MFC GABA+ in psychosis.

### 4.1 Increased GABA+ concentrations in the rostral MFC in early psychosis

The first process is distinguished by elevated GABA+ concentrations in the rostral MFC, which occur early in the illness and may be driven down by treatment or other factors which has led to contradictory findings in the literature. This process may reflect a primary pathophysiological process or a compensatory response – for example, to environmental stressors. A chronic unpredictable stress paradigm in rodents has been shown to increase ACC GABA levels, as measured by MRS (Perrine et al., 2014). Alternatively, increased GABA may be a secondary response to glutamatergic dysfunction. For example, ketamine infusion, which blocks excitatory NMDA receptors, has been shown to increase medial prefrontal cortex GABA levels in humans (Rodriguez et al., 2015). We examined Glx concentrations in our sample and found no evidence for abnormal concentrations in psychosis. Glx, however, is the combined signal of glutamate and its precursor glutamine, and a balanced cycling between these two amino acids is essential for normal brain functioning. Investigating Glx concentrations may miss subtle shifts in the balances between glutamate (Glu) and glutamine (Gln), and therefore fail to detect glutamatergic dysfunction. A recent meta-analysis of glutamatergic metabolites does provide some evidence for elevated Glx and Glu in treatment resistant schizophrenia (Nakahara et al., 2021), lending support to the hypothesis of glutamatergic dysfunction in psychosis.

When comparing the APS group with controls, we found a trend suggesting increased rostral MFC GABA+ in APS, controlling for levels of negative affect. While this effect did not reach significance and should be interpreted with caution, our APS sample was small, and this analysis was undoubtedly underpowered. This trend is in line with previous evidence indicating elevated rostral MFC GABA+ in individuals at an ultra-high risk of developing psychosis (De La Fuente-Sandoval et al., 2015), and indicates that processes resulting in abnormal GABA concentrations in the rostral MFC may be present prior to the conversion to full psychosis.

### 4.2 Increased Negative Affect is associated with reduced GABA+ concentrations

As we note, multiple factors may drive GABA+ levels down and obscure group differences, and we propose there is evidence for a second process in which increased NA is associated with reduced rostral MFC GABA+, as well as the altered BOLD response observed in benzodiazepine challenge studies published by our group (Taylor et al., 2014; Tso et al., 2015). In the current study, we observed significantly higher levels of NA in our FEP group when compared with controls, and once we controlled for levels of NA in our statistical analysis, we found significant group differences in rostral MFC GABA+ concentrations. These differences were at a trend level without the inclusion of levels of NA in our model, indicating that NA may have been driving GABA+ levels down and obscuring group differences. The existence of two processes affecting GABA+ levels could explain the otherwise confusing finding from de la Fuente-Sandoval and colleagues, who found an inverse correlation between rostral MFC GABA levels and PANSS general psychopathology scores – which also capture NA - in first episode psychosis patients, who otherwise had higher GABA levels (de la Fuente-Sandoval et al., 2018).

Evidence from benzodiazepine challenge studies has linked greater levels of NA with altered GABAergic function in the dorsomedial prefrontal cortex while viewing salient stimuli (Taylor et al., 2014) and in the visual cortex during a face processing task (Tso et al., 2015). These findings indicate that this process may be more widespread throughout the cortex rather than confined to frontal regions, although we found no evidence for a relationship with NA in our midline occipital MRS voxel. In addition, Taylor and colleagues (2014) found that negative affect correlated positively with the altered BOLD signal in both patients and controls, suggesting that this process may be present on a continuum from psychosis to disease.

Reviewing the literature provides further support for a link between reduced GABA and NA. Considerable evidence has supported the involvement of reduced GABAergic activity in patients with major depressive disorder (MDD), who have an abundance of NA (Duman et al., 2019). GABA MRS studies have shown reduced GABA in MDD patients, including the MFC where we placed our voxels (Gabbey et al., 2013; Kantrowitz et al., 2021) .The demonstration that neuropeptides which potentiate GABAergic transmission have rapid antidepressant effects provides additional evidence of a role for GABA in NA (Fee et al., 2017) in experimental paradigms, inducing NA with a fear-of shock challenge acutely decreased rostral MFC GABA in healthy controls (Hasler et al., 2010). GAD1 (the gene coding for the GAD67 protein) knock-down mouse models have demonstrated impaired fear extinction (Brown et al., 2015) and emotion regulation but preserved working memory function (Kolata et al., 2018). Although as the major inhibitory neurotransmitter, GABA activity is tightly coupled in an excitatory-inhibitory balance which is difficult to easily characterize in pathological states, multiple lines of evidence show signs of reduced GABA activity, linked to various NA states.

### 4.3 Limitations and Future Directions

A key limitation of this study was its modest sample size, which means the results we describe should be interpreted with the appropriate caution, and replication of these effects is vital. As we note above, the small sample of participants with APS perhaps meant that our exploration of GABA+ in these individuals was underpowered. In addition, the majority of our FEP group and one member of our APS group were receiving anti-psychotic medication, which meant our sample was not free of medication confounds. Since our sample size was relatively small, we lacked the statistical power to probe the influence of medication on GABA+ concentrations while investigating linkages between GABA+ and negative affect. Prior research indicates that the relationship between GABA+ concentrations and anti-psychotic medications is perhaps complex, with evidence showing a negative correlation between GABA+ levels in the ACC and anti-psychotic dose (Tayoshi et al., 2010) and reductions in mPFC GABA levels following a 4 week course of atypical anti-psychotics (de la Fuente-Sandoval et al., 2018), contrasted with findings that a 6 month treatment with atypical anti-psychotics did not affect GABAergic transmission in the frontal lobe, left basal ganglia or parietal-occipital lobe (Goto et al., 2010). Further investigations of linkages between GABA concentrations and negative affect would benefit from a larger sample size which allows the impact of medication to be carefully explored. Additionally, future work should extend our MRS findings, and the findings from our group’s pharmacological probe of the impact of lorazepam on the BOLD response in the MFC, to investigate if reduced GABA levels in the rostral MFC are associated with an altered BOLD response in the same individuals.

### 4.4 Conclusions

In conclusion, we found evidence for significantly elevated rostral MFC GABA+ concentrations in psychosis, when controlling for levels of negative affect. When considered alongside previously published demonstrations of altered BOLD signal in response to a benzodiazepine challenge, as well as linkages between GABA and negative affective states, we suggest the presence of two pathological processes involving GABA. The first is distinct from negative affect and is characterized by an increase of GABA in the rostral MFC which occurs early in the illness. The second process is reflected by reduced GABA which is associated with increased negative affect and an altered BOLD response to benzodiazepine. While more data is required to untangle these processes, and clarify GABAergic dysfunction in the psychosis spectrum, NA is a strong, transdiagnostic predictor of functional outcome, and these findings establish potential therapeutic leverage points linking GABAergic treatments, with NA as the clinical outcome.

## Data Availability

All data produced in the present study are available upon reasonable request to the authors

## REFERENCES

Behrens, M. M., Ali, S. S., Dao, D. N., Lucero, J., Shekhtman, G., Quick, K. L., & Dugan, L. L. (2007). Ketamine-induced loss of phenotype of fast-spiking interneurons is mediated by NADPH-oxidase. Science, 318(5856), 1645–1647. https://doi.org/10.1126/SCIENCE.1148045

Blanchard, J. J., Mueser, K. T., & Bellack, A. S. (1998). Anhedonia, positive and negative affect, and social functioning in schizophrenia. Schizophrenia Bulletin, 24(3), 413–424. https://doi.org/10.1093/oxfordjournals.schbul.a033336

Brown, J. A., Ramikie, T. S., Schmidt, M. J., Báldi, R., Garbett, K., Everheart, M. G., Warren, L. E., Gellért, L., Horváth, S., Patel, S., & Mirnics, K. (2015). Inhibition of parvalbumin-expressing interneurons results in complex behavioral changes. Molecular Psychiatry, 20(12), 1499–1507. https://doi.org/10.1038/MP.2014.192

Burbridge, J. A., & Barch, D. M. (2002). Emotional valence and reference disturbance in schizophrenia. Journal of Abnormal Psychology, 111(1), 186–191. https://doi.org/10.1037/0021-843X.111.1.186

Chen, C. M. A., Stanford, A. D., Mao, X., Abi-Dargham, A., Shungu, D. C., Lisanby, S. H., Schroeder, C. E., & Kegeles, L. S. (2014). GABA level, gamma oscillation, and working memory performance in schizophrenia. NeuroImage: Clinical, 4, 531–539. https://doi.org/10.1016/J.NICL.2014.03.007

Chen, T., Wang, Y., Zhang, J., Wang, Z., Xu, J., Li, Y., Yang, Z., & Liu, D. (2017). Abnormal Concentration of GABA and Glutamate in The Prefrontal Cortex in Schizophrenia.-An in Vivo 1H-MRS Study. Shanghai Archives of Psychiatry, 29(5), 277–286. https://doi.org/10.11919/j.issn.1002-0829.217004

Chiu, P. W., Lui, S. S. Y., Hung, K. S. Y., Chan, R. C. K., Chan, Q., Sham, P. C., Cheung, E. F. C., & Mak, H. K. F. (2018). In vivo gamma-aminobutyric acid and glutamate levels in people with first-episode schizophrenia: A proton magnetic resonance spectroscopy study. Schizophrenia Research, 193, 295–303. https://doi.org/10.1016/j.schres.2017.07.021

Cohen, A. S., & Minor, K. S. (2010). Emotional experience in patients with schizophrenia revisited: Meta-analysis of laboratory studies. Schizophrenia Bulletin, 36(1), 143–150. https://doi.org/10.1093/schbul/sbn061

Cohen, S., Kamarck, T., & Mermelstein, R. (1983). Perceived Stress Scale. In Journal of Health and Social Behavior.

Curley, A. A., Arion, D., Volk, D. W., Asafu-Adjei, J. K., Sampson, A. R., Fish, K. N., & Lewis, D. A. (2011). Cortical deficits of glutamic acid decarboxylase 67 expression in schizophrenia: Clinical, protein, and cell type-specific features. The American Journal of Psychiatry, 168(9), 921–929. https://doi.org/10.1176/APPI.AJP.2011.11010052

de la Fuente-Sandoval, C., Reyes-Madrigal, F., Mao, X., León-Ortiz, P., Rodríguez-Mayoral, O., Jung-Cook, H., Solís-Vivanco, R., Graff-Guerrero, A., & Shungu, D. C. (2018). Prefrontal and Striatal Gamma-Aminobutyric Acid Levels and the Effect of Antipsychotic Treatment in First-Episode Psychosis Patients. Biological Psychiatry, 83(6), 475–483. https://doi.org/10.1016/j.biopsych.2017.09.028

De La Fuente-Sandoval, C., Reyes-Madrigal, F., Mao, X., León-Ortiz, P., Rodríguez-Mayoral, O., Solís-Vivanco, R., Favila, R., Graff-Guerrero, A., & Shungu, D. C. (2015). Cortico-striatal GABAergic and glutamatergic dysregulations in subjects at ultra-high risk for psychosis investigated with proton magnetic resonance spectroscopy. International Journal of Neuropsychopharmacology, 19(3), 1–10. https://doi.org/10.1093/ijnp/pyv105

Docherty, N. M. (1996). Affective reactivity of symptoms as a process discriminator in schizophrenia. In Journal of Nervous and Mental Disease (Vol. 184, Issue 9, pp. 535–541). J Nerv Ment Dis. https://doi.org/10.1097/00005053-199609000-00004

Duman, R. S., Sanacora, G., & Krystal, J. H. (2019). Altered Connectivity in Depression: GABA and Glutamate Neurotransmitter Deficits and Reversal by Novel Treatments. Neuron, 102(1), 75–90. https://doi.org/10.1016/J.NEURON.2019.03.013

Edden, R. A. E., Puts, N. A. J., Harris, A. D., Barker, P. B., & Evans, C. J. (2014). Gannet: A batch-processing tool for the quantitative analysis of gamma-aminobutyric acid-edited MR spectroscopy spectra. Journal of Magnetic Resonance Imaging, 40(6), 1445–1452. https://doi.org/10.1002/jmri.24478

Fee, C., Banasr, M., & Sibille, E. (2017). Somatostatin-positive GABA Interneuron Deficits in Depression: Cortical Microcircuit and Therapeutic Perspectives. Biological Psychiatry, 82(8), 549. https://doi.org/10.1016/J.BIOPSYCH.2017.05.024

First, M. B., Spitzer, R., Gibbon, M., & Williams, J. (1996). Structured Clinical Interview for Axis I Disorders—Patient Edition. Biometrics Research, New York State Psychiatric Institute.

Ganji, S. K., An, Z., Banerjee, A., Madan, A., Hulsey, K. M., & Choi, C. (2014). Measurement of regional variation of GABA in the human brain by optimized point-resolved spectroscopy at 7 T in vivo. NMR in Biomedicine, 27(10), 1167–1175. https://doi.org/10.1002/NBM.3170

Gabbay, V., Mao, X., Klein, R. G., Ely, B. A., Babb, J. S., Panzer, A. M., & Shungu, D. C. (2012). Anterior cingulate cortex γ-aminobutyric acid in depressed adolescents: Relationship to anhedonia. Archives of general psychiatry, 69(2), 139–149.

Goto, N., Yoshimura, R., Kakeda, S., Moriya, J., Hori, H., Hayashi, K., Ikenouchi-Sugita, A., Nakano-Umene, W., Katsuki, A., Nishimura, J., Korogi, Y., & Nakamura, J. (2010). No alterations of brain GABA after 6 months of treatment with atypical antipsychotic drugs in early-stage first-episode schizophrenia. Progress in Neuro-Psychopharmacology and Biological Psychiatry, 34(8), 1480–1483. https://doi.org/10.1016/j.pnpbp.2010.08.007

Goto, N., Yoshimura, R., Moriya, J., Kakeda, S., Ueda, N., Ikenouchi-Sugita, A., Umene-Nakano, W., Hayashi, K., Oonari, N., Korogi, Y., & Nakamura, J. (2009). Reduction of brain γ-aminobutyric acid (GABA) concentrations in early-stage schizophrenia patients: 3T Proton MRS study. In Schizophrenia Research (Vol. 112, Issues 1–3, pp. 192–193). https://doi.org/10.1016/j.schres.2009.04.026

Grove, T. B., Tso, I. F., Chun, J., Mueller, S. A., Taylor, S. F., Ellingrod, V. L., McInnis, M. G., & Deldin, P. J. (2016). Negative affect predicts social functioning across schizophrenia and bipolar disorder: Findings from an integrated data analysis. Psychiatry Research, 243, 198–206. https://doi.org/10.1016/j.psychres.2016.06.031

Guidotti, A., Auta, J., Davis, J. M., Gerevini, V. D., Dwivedi, Y., Grayson, D. R., Impagnatiello, F., Pandey, G., Pesold, C., Sharma, R., Uzunov, D., & Costa, E. (2000). Decrease in Reelin and Glutamic Acid Decarboxylase67 (GAD67) Expression in Schizophrenia and Bipolar Disorder: A Postmortem Brain Study. Archives of General Psychiatry, 57(11), 1061–1069. https://doi.org/10.1001/ARCHPSYC.57.11.1061

Harris, A. D., Puts, N. A. J., & Edden, R. A. E. (2015). Tissue correction for GABA-edited MRS: Considerations of voxel composition, tissue segmentation, and tissue relaxations. Journal of Magnetic Resonance Imaging, 42(5), 1431–1440. https://doi.org/10.1002/jmri.24903

Hasler, G., Van Der Veen, J. W., Grillon, C., Drevets, W. C., & Shen, J. (2010). Effect of Acute Psychological Stress on Prefrontal GABA Concentration Determined by Proton Magnetic Resonance Spectroscopy. The American Journal of Psychiatry, 167(10), 1226. https://doi.org/10.1176/APPI.AJP.2010.09070994

Holt, D. J., Titone, D., Long, L. S., Goff, D. C., Cather, C., Rauch, S. L., Judge, A., & Kuperberg, G. R. (2006). The misattribution of salience in delusional patients with schizophrenia. Schizophrenia Research, 83(2–3), 247–256. https://doi.org/10.1016/j.schres.2005.12.858

Horan, W. P., Blanchard, J. J., Clark, L. A., & Green, M. F. (2008). Affective traits in schizophrenia and schizotypy. In Schizophrenia Bulletin (Vol. 34, Issue 5, pp. 856–874). https://doi.org/10.1093/schbul/sbn083

Horan, W. P., Ventura, J., Nuechterlein, K. H., Subotnik, K. L., Hwang, S. S., & Mintz, J. (2005). Stressful life events in recent-onset schizophrenia: Reduced frequencies and altered subjective appraisals. Schizophrenia Research, 75(2–3), 363–374. https://doi.org/10.1016/j.schres.2004.07.019

Jones, S. R., & Fernyhough, C. (2007). A new look at the neural diathesis-stress model of schizophrenia: The primacy of social-evaluative and uncontrollable situations. Schizophrenia Bulletin, 33(5), 1171–1177. https://doi.org/10.1093/schbul/sbl058

Kantrowitz, J. T., Dong, Z., Milak, M. S., Rashid, R., Kegeles, L. S., Javitt, D. C., & John Mann, J. (2021). Ventromedial prefrontal cortex/anterior cingulate cortex Glx, glutamate, and GABA levels in medication-free major depressive disorder. Translational psychiatry, 11(1), 1–6.

Kegeles, L. S., Mao, X., Stanford, A. D., Girgis, R., Ojeil, N., Xu, X., Gil, R., Slifstein, M., Abi-Dargham, A., Lisanby, S. H., & Shungu, D. C. (2012). Elevated prefrontal cortex γ-aminobutyric acid and glutamate-glutamine levels in schizophrenia measured in vivo with proton magnetic resonance spectroscopy. Archives of General Psychiatry, 69(5), 449–459. https://doi.org/10.1001/archgenpsychiatry.2011.1519

Kolata, S. M., Nakao, K., Jeevakumar, V., Farmer-Alroth, E. L., Fujita, Y., Bartley, A. F., Jiang, S. Z., Rompala, G. R., Sorge, R. E., Jimenez, D. V., Martinowich, K., Mateo, Y., Hashimoto, K., Dobrunz, L. E., & Nakazawa, K. (2018). Neuropsychiatric Phenotypes Produced by GABA Reduction in Mouse Cortex and Hippocampus. Neuropsychopharmacology, 43(6), 1445. https://doi.org/10.1038/NPP.2017.296

Lewis, D. A., Curley, A. A., Glausier, J. R., & Volk, D. W. (2012). Cortical parvalbumin interneurons and cognitive dysfunction in schizophrenia. Trends in Neurosciences, 35(1), 57–67. https://doi.org/10.1016/J.TINS.2011.10.004

Lin, A., Andronesi, O., Bogner, W., Choi, I. Y., Coello, E., Cudalbu, C., Juchem, C., Kemp, G. J., Kreis, R., Krššák, M., Lee, P., Maudsley, A. A., Meyerspeer, M., Mlynarik, V., Near, J., Öz, G., Peek, A. L., Puts, N. A., Ratai, E. M., Mullins, P. G. (2021). Minimum Reporting Standards for in vivo Magnetic Resonance Spectroscopy (MRSinMRS): Experts’ consensus recommendations. NMR in Biomedicine, 34(5), e4484. https://doi.org/10.1002/NBM.4484

Liu, W. S., Pesold, C., Rodriguez, M. A., Carboni, G., Auta, J., Lacor, P., Larson, J., Condie, B. G., Guidotti, A., & Costa, E. (2001). Down-regulation of dendritic spine and glutamic acid decarboxylase 67 expressions in the reelin haploinsufficient heterozygous reeler mouse. Proceedings of the National Academy of Sciences of the United States of America, 98(6), 3477–3482. https://doi.org/10.1073/PNAS.051614698

Liu, Y., Ouyang, P., Zheng, Y., Mi, L., Zhao, J., Ning, Y., & Guo, W. (2021). A Selective Review of the Excitatory-Inhibitory Imbalance in Schizophrenia: Underlying Biology, Genetics, Microcircuits, and Symptoms. Frontiers in cell and developmental biology, 9, 664535. https://doi.org/10.3389/fcell.2021.664535

Menschikov, P. E., Semenova, N. A., Ublinskiy, M. V., Akhadov, T. A., Keshishyan, R. A., Lebedeva, I. S., Omelchenko, M. A., Kaleda, V. G., & Varfolomeev, S. D. (2016). 1H-MRS and MEGA-PRESS pulse sequence in the study of balance of inhibitory and excitatory neurotransmitters in the human brain of ultra-high risk of schizophrenia patients. Doklady Biochemistry and Biophysics, 468(1), 168–172. https://doi.org/10.1134/S1607672916030029

Mescher, M., Merkle, H., Kirsch, J., Garwood, M., & Gruetter, R. (1998). Simultaneous in vivo spectral editing and water suppression. NMR in Biomedicine, 11(6), 266–272. https://doi.org/10.1002/(SICI)1099-1492(199810)11:6<266::AID-NBM530>3.0.CO;2-J

Miller, T. J., McGlashan, T. H., Rosen, J. L., Cadenhead, K., Ventura, J., McFarlane, W., Perkins, D. O., Pearlson, G. D., & Woods, S. W. (2003). Prodromal assessment with the structured interview for prodromal syndromes and the scale of prodromal symptoms: Predictive validity, interrater reliability, and training to reliability. Schizophrenia Bulletin, 29(4), 703–715. https://doi.org/10.1093/OXFORDJOURNALS.SCHBUL.A007040

Mullins, P. G., McGonigle, D. J., O’Gorman, R. L., Puts, N. A. J., Vidyasagar, R., Evans, C. J., Edden, R. A. E., Brookes, M. J., Garcia, A., Foerster, B. R., Petrou, M., Price, D., Solanky, B. S., Violante, I. R., Williams, S., & Wilson, M. (2014). Current practice in the use of MEGA-PRESS spectroscopy for the detection of GABA. In NeuroImage (Vol. 86, pp. 43–52). Academic Press. https://doi.org/10.1016/j.neuroimage.2012.12.004

Myin-Germeys, I., Delespaul, P. A. E. G., & DeVries, M. W. (2000). Schizophrenia patients are more emotionally active than is assumed based on their behavior. Schizophrenia Bulletin, 26(4), 847–854. https://doi.org/10.1093/oxfordjournals.schbul.a033499

Myin-Germeys, I., & van Os, J. (2007). Stress-reactivity in psychosis: Evidence for an affective pathway to psychosis. Clinical Psychology Review, 27(4), 409–424. https://doi.org/10.1016/J.CPR.2006.09.005

Nakahara, T., Tsugawa, S., Noda, Y., Ueno, F., Honda, S., Kinjo, M., Segawa, H., Hondo, N., Mori, Y., Watanabe, H., Nakahara, K., Yoshida, K., Wada, M., Tarumi, R., Iwata, Y., Plitman, E., Moriguchi, S., de la Fuente-Sandoval, C., Uchida, H., … Nakajima, S. (2021). Glutamatergic and GABAergic metabolite levels in schizophrenia-spectrum disorders: A meta-analysis of 1H-magnetic resonance spectroscopy studies. In Molecular Psychiatry. https://doi.org/10.1038/s41380-021-01297-6

Öngür, D., Prescot, A. P., Jensen, J. E., Cohen, B. M., & Renshaw, P. F. (2009). CREATINE ABNORMALITIES IN SCHIZOPHRENIA AND BIPOLAR DISORDER. Psychiatry Research, 172(1), 44. https://doi.org/10.1016/J.PSCYCHRESNS.2008.06.002

Öngür, D., Prescot, A. P., McCarthy, J., Cohen, B. M., & Renshaw, P. F. (2010). Elevated gamma-aminobutyric acid levels in chronic schizophrenia. Biological Psychiatry, 68(7), 667–670. https://doi.org/10.1016/j.biopsych.2010.05.016

Perrine, S. A., Ghoddoussi, F., Michaels, M. S., Sheikh, I. S., McKelvey, G., & Galloway, M. P. (2014). Ketamine reverses stress-induced depression-like behavior and increased GABA levels in the anterior cingulate: An 11.7T 1H-MRS study in rats. Progress in Neuro-Psychopharmacology and Biological Psychiatry, 51, 9–15. https://doi.org/10.1016/j.pnpbp.2013.11.003

Phillips, L. J., Francey, S. M., Edwards, J., & McMurray, N. (2007). Stress and psychosis: Towards the development of new models of investigation. Clinical Psychology Review, 27(3), 307–317. https://doi.org/10.1016/J.CPR.2006.10.003

Rodriguez, C. I., Kegeles, L. S., Levinson, A., Ogden, R. T., Mao, X., Milak, M. S., Vermes, D., Xie, S., Hunter, L., Flood, P., Moore, H., Shungu, D. C., & Simpson, H. B. (2015). In vivo effects of ketamine on glutamate-glutamine and gamma-aminobutyric acid in obsessive-compulsive disorder: Proof of concept. Psychiatry Research - Neuroimaging, 233(2), 141–147. https://doi.org/10.1016/j.pscychresns.2015.06.001

Rowland, L. M., Kontson, K., West, J., Edden, R. A., Zhu, H., Wijtenburg, S. A., Holcomb, H. H., & Barker, P. B. (2013). In vivo measurements of glutamate, GABA, and NAAG in schizophrenia. Schizophrenia Bulletin, 39(5), 1096–1104. https://doi.org/10.1093/schbul/sbs092

Stan, A. D., Schirda, C. V., Bertocci, M. A., Bebko, G. M., Kronhaus, D. M., Aslam, H. A., LaBarbara, E. J., Tanase, C., Lockovich, J. C., Pollock, M. H., Stiffler, R. S., & Phillips, M. L. (2014). Glutamate and GABA contributions to medial prefrontal cortical activity to emotion: implications for mood disorders. Psychiatry research, 223(3), 253–260. https://doi.org/10.1016/j.pscychresns.2014.05.016

Schmidt, M. J., & Mirnics, K. (2015). Neurodevelopment, GABA system dysfunction, and schizophrenia. NeuropsychopharmacologyfI: Official Publication of the American College of Neuropsychopharmacology, 40(1), 190–206. https://doi.org/10.1038/NPP.2014.95

Takahashi, T., & Suzuki, M. (2018). Brain morphologic changes in early stages of psychosis: Implications for clinical application and early intervention. Psychiatry and Clinical Neurosciences, 72(8), 556–571. https://doi.org/10.1111/PCN.12670

Takao, K., Kobayashi, K., Hagihara, H., Ohira, K., Shoji, H., Hattori, S., Koshimizu, H., Umemori, J., Toyama, K., Nakamura, H. K., Kuroiwa, M., Maeda, J., Atsuzawa, K., Esaki, K., Yamaguchi, S., Furuya, S., Takagi, T., Walton, N. M., Hayashi, N., … Miyakawa, T. (2013). Deficiency of Schnurri-2, an MHC Enhancer Binding Protein, Induces Mild Chronic Inflammation in the Brain and Confers Molecular, Neuronal, and Behavioral Phenotypes Related to Schizophrenia. Neuropsychopharmacology 2013 38:8, 38(8), 1409–1425. https://doi.org/10.1038/npp.2013.38

Taylor, S. F., Demeter, E., Phan, K. L., Tso, I. F., & Welsh, R. C. (2014). Abnormal GABAergic Function and Negative Affect in Schizophrenia. Neuropsychopharmacology, 39(4), 1000–1008. https://doi.org/10.1038/npp.2013.300

Tayoshi, S., Nakataki, M., Sumitani, S., Taniguchi, K., Shibuya-Tayoshi, S., Numata, S. Iga, J. ichi, Ueno, S. ichi, Harada, M., & Ohmori, T. (2010). GABA concentration in schizophrenia patients and the effects of antipsychotic medication: A proton magnetic resonance spectroscopy study. Schizophrenia Research, 117(1), 83–91. https://doi.org/10.1016/j.schres.2009.11.011

Thakkar, K. N., Rösler, L., Wijnen, J. P., Boer, V. O., Klomp, D. W. J., Cahn, W., Kahn, R. S., & Neggers, S. F. W. (2017). 7T Proton Magnetic Resonance Spectroscopy of Gamma-Aminobutyric Acid, Glutamate, and Glutamine Reveals Altered Concentrations in Patients With Schizophrenia and Healthy Siblings. Biological Psychiatry, 81(6), 525–535. https://doi.org/10.1016/j.biopsych.2016.04.007

Torrey, E. F., Barci, B. M., Webster, M. J., Bartko, J. J., Meador-Woodruff, J. H., & Knable, M. B. (2005). Neurochemical markers for schizophrenia, bipolar disorder, and major depression in postmortem brains. 57(3), 252–260. https://doi.org/10.1016/J.BIOPSYCH.2004.10.019

Tso, I. F., Fang, Y., Phan, K. L., Welsh, R. C., & Taylor, S. F. (2015). Abnormal GABAergic function and face processing in schizophrenia: A pharmacologic-fMRI study. Schizophrenia Research, 168(1–2), 338–344. https://doi.org/10.1016/j.schres.2015.08.022

Tso, I. F., Grove, T. B., & Taylor, S. F. (2012). Self-assessment of psychological stress in schizophrenia: Preliminary evidence of reliability and validity. Psychiatry Research, 195(1–2), 39–44. https://doi.org/10.1016/j.psychres.2011.07.009

Wang, J. J., Wang, J. J., Tang, Y., Zhang, T., Cui, H., Xu, L., Zeng, B., Li, Y., Li, G., Li, C., Liu, H., Lu, Z., & Zhang, J. (2016). Reduced γ-Aminobutyric Acid and Glutamate+Glutamine Levels in Drug-Naïve Patients with First-Episode Schizophrenia but Not in Those at Ultrahigh Risk. Neural Plasticity, 2016. https://doi.org/10.1155/2016/3915703

Woo, T. U. W., Kim, A. M., & Viscidi, E. (2008). Disease-specific alterations in glutamatergic neurotransmission on inhibitory interneurons in the prefrontal cortex in schizophrenia. 1218, 267–277. https://doi.org/10.1016/J.BRAINRES.2008.03.092

Yang, Z., Zhu, Y., Song, Z., Mei, L., Zhang, J., Chen, T., Wang, Y., Xu, Y., Jiang, K., Li, Y., & Liu, D. (2015). Comparison of the density of gamma-aminobutyric acid in the ventromedial prefrontal cortex of patients with first-episode psychosis and healthy controls. Shanghai Archives of Psychiatry, 27(6), 341–347. https://doi.org/10.11919/j.issn.1002-0829.215130

Yoon, J. H., Maddock, R. J., Rokem, A., Silver, M. A., Minzenberg, M. J., Ragland, J. D., & Carter, C. S. (2010). GABA Concentration Is Reduced in Visual Cortex in Schizophrenia and Correlates with Orientation-Specific Surround Suppression. Journal of Neuroscience, 30(10), 3777–3781. https://doi.org/10.1523/JNEUROSCI.6158-09.2010

