## Supplemental materials for "Increased rostral medial frontal GABA+ in early psychosis is obscured by levels of negative affect"

| Supplemental Table 1: MRS data spectral quality measures | | | | | |
| --- | --- | --- | --- | --- | --- |
|  |  | Group | | | Group difference |
| Voxel | Measure | HC | APS | FEP |  |
| MFC | GABA+ FWHM | 20.10 (±3.99) | 21.15 (±3.83) | 20.17 (±4.25) | F_2,28_ = 0.17, *p* = .84 |
|  | GABA+ SNR | 6.87 (±1.17) | 6.29 (±1.81) | 7.21 (±2.13) | F_2,28_ = 0.63, *p* = .54 |
|  | Glx FWHM | 16.04 (±2.50) | 15.94 (±2.15) | 15.69 (±1.26) | F_2,32_ = 0.10, *p* = .90 |
|  | Glx SNR | 18.05 (±4.56) | 17.67 (±5.16) | 15.85 (±3.44) | F_2,32_ = 0.97, *p* = .39 |
| Occ | GABA+ FWHM | 16.26 (±2.01) | 17.55 (±2.34) | 16.84 (±2.17) | F_2,32_ = 0.82, *p* = .45 |
|  | GABA+ SNR | 15.47 (±3.20) | 16.07 (±2.81) | 14.60 (±3.31) | F_2,32_ = 0.52, *p* = .60 |
|  | Glx FWHM | 15.93 (±2.07) | 16.83 (±1.51) | 15.23 (±0.94) | F_2,32_ = 2.16, *p* = .13 |
|  | Glx SNR | 16.32 (±3.51) | 15.88 (±1.98) | 15.38 (±2.99) | F_2,32_ = 0.33, *p* = .72 |

| Supplemental Table 2: Uncorrected metabolite levels in rostral MFC and midline occipital cortex | | | | | | |
| --- | --- | --- | --- | --- | --- | --- |
|  | | Group | | | Group difference | Group difference, corrected for PSI |
| Voxel | Measure | HC | APS | FEP |  |  |
| MFC | GABA+/Cr | 0.06 (±0.01) | 0.06 (±0.01) | 0.07 (±0.02) | F_2,28_ = 0.83, *p* = .45 | F_2,27_ = 2.36, *p* = .10 |
|  | GABA+/H2O | 1.29 (±0.25) | 1.23 (±0.01) | 1.49 (±0.31) | F_2,28_ = 2.69, *p* = .09 | F_2,27_ = 3.51, *p* = .04 |
|  | Glx/Cr | 0.12 (±0.01) | 0.12 (±0.02) | 0.12 (±0.01) | F_2,28_ = 0.24, *p* = .79 | F_2,31_ = 0.79, *p* = .46 |
|  | Glx/H2O | 8.75 (±1.12) | 8.54 (±0.92) | 8.57 (±2.32) | F_2,28_ = 0.06, *p* = .94 | F_2,31_ = 0.28, *p* = .76 |
| Occ | GABA+/Cr | 0.10 (±0.01) | 0.10 (±0.02) | 0.10 (±0.01) | F_2,32_ = 0.58, *p* = .56 | F_2,31_ = 0.57, *p* = .57 |
|  | GABA+/H2O | 2.16 (±0.31) | 2.30 (±0.32) | 2.12 (±0.32) | F_2,32_ = 0.72, *p* = .50 | F_2,31_ = 0.66, *p* = .52 |
|  | Glx/Cr | 0.09 (±0.01) | 0.09 (±0.01) | 0.09 (±0.02) | F_2,32_ = 0.35, *p* = .71 | F_2,32_ = 1.11, *p* = .34 |
|  | Glx/H2O | 7.48 (±1.07) | 7.45 (±0.95) | 6.94 (±1.14) | F_2,32_ = 1.01, *p* = .38 | F_2,32_ = 1.54, *p* = .23 |

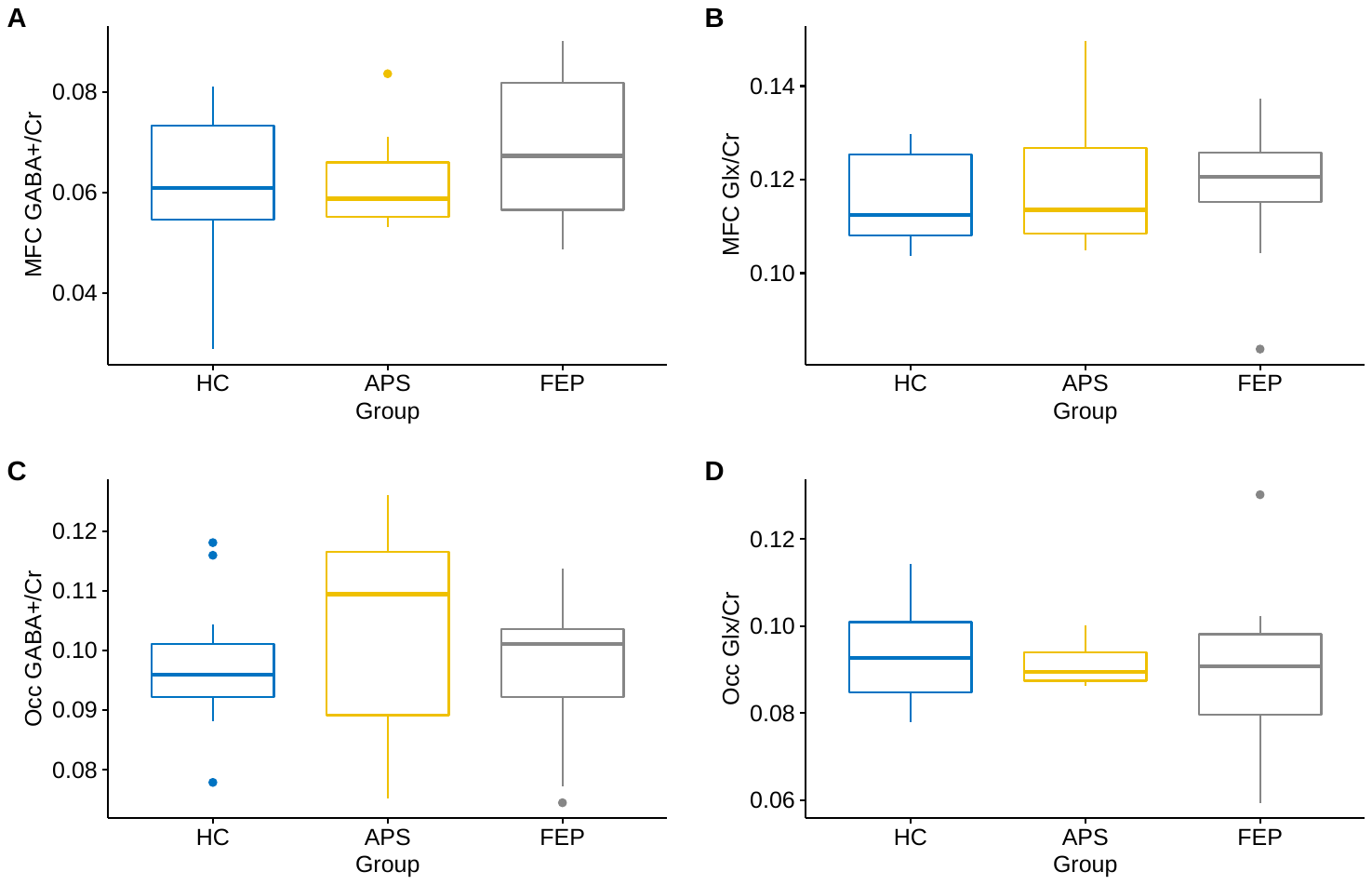

Supplemental Figure 1: Boxplots demonstrating rostral MFC (A) GABA+ and (B) Glx and midline occipital (C) GABA+ and (D) Glx when concentrations are not tissue corrected and are referenced to Cr.

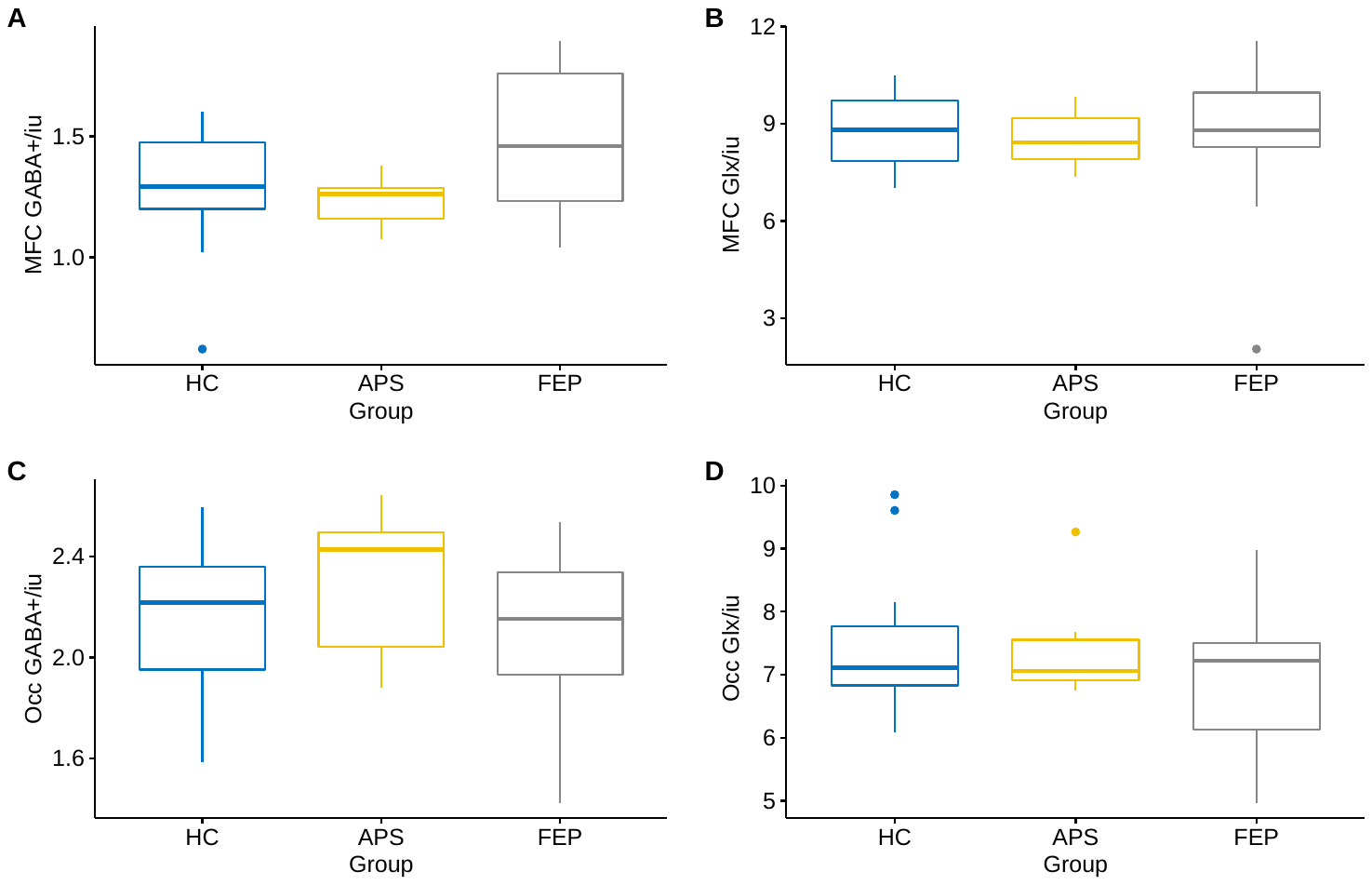

Supplemental Figure 2: Boxplots demonstrating rostral MFC (A) GABA+ and (B) Glx and midline occipital (C) GABA+ and (D) Glx when concentrations are not tissue corrected and are referenced to H2O

| Supplemental Table 3: Minimum Reporting Standard for in-vivo Magnetic Resonance Spectroscopy: Experts Consensus Recommendation | |
| --- | --- |
| 1. **Hardware** | |
| a. Field Strength [T] | 3T |
| b. Manufacturer | Philips |
| c. Model (software version if available) | Ingenia SW R5.3 |
| d. RF coils: nuclei (transmit/receive), number of channels, type, body part | 32 channel head coil |
| e. Additional hardware |  |
| 2. **Acquisition** |  |
| a. Pulse sequence | MEGA-PRESS (Mescher et al., 1998) |
| b. Volume of Interest (VOI) locations | Rostral medial frontal cortex (rMFC) and occipital cortex (Occ)  Figure 1 presents example placement, Figure 2 presents heatmaps demonstrating VOI overlap across all participants |
| c. Nominal VOI size [cm3, mm3] | rMFC: 34 x 36 x 25 mm  Occ: 35 x 25 x 25 mm |
| d. Repetition time (*T*_R_), echo time (*T*_E_) [ms, s] | T_R_ = 2000ms, T_E_ = 68 ms (  TE1 = 15ms, T_E_2 = 53ms) |
| e. Total number of excitations of or acquisitions per spectrum   In time series for kinetic studies   i. Number of averaged spectra (NA) per time point ii. Averaging method (eg block-wise or moving average) iii. Total number of spectra (acquired/in time series) | 256 (128 ON interleaved with 128 OFF) |
| f. Additional sequence parameters (spectral width in Hz, number of spectral points, frequency offsets)   If STEAM: mixing time (*T*_M_)  If MRSI: 2D or 3D, FOX in all directions, matrix size, acceleration factors, sampling method | Spectral width = 2 kHz  Data points = 2000  Frequency selective editing pulses applied at:  ON pulse = 1.9 ppm  OFF pulse = 7.46 ppm |
| g. Water suppression method |  |
| h. Shimming method, reference peak, and thresholds for “acceptance of shim” chosen |  |
| i. Triggering or motion correction method (respiratory, peripheral, cardiac triggering, incl. device used and delays | None |
| 3. **Data analysis methods and outputs** |  |
| a. Analysis software | Gannet 3.1 |
| b. Processing steps deviating from quoted reference or product | No deviation |
| c. Output measure (eg absolute concentration, institutional units, ratio), processing steps deviating from quote reference or product | In main manuscript: GABA+/H_2_O α-corrected and Glx/H_2_O α-corrected  In supplemental Table 2: GABA+/H_2_O, Glx/H_2_O, GABA+/Cr, Glx/Cr |
| d. Quantification references and assumptions, fitting model assumption |  |
| 4. **Data quality** |  |
| a. Reported variables (SNR, linewidth (with reference peaks)) | Linewidth and SNR reported in Supplemental Table 1 |
| b. Data exclusion criteria | Data were excluded if Fit Error (%) was greater than 15, or if metabolite estimate was a large outlier (more than ± 5SDs from mean) |
| c. Quality measures of post-processing model fitting (eg CRLB, goodness of fit, SD of residual) |  |
| d. Sample spectrum | Presented in main manuscript, Figure 1 |
